# Retrospective analysis of clinical and environmental genotyping reveals persistence of *Pseudomonas aeruginosa* in the water system of a large tertiary children’s hospital in England

**DOI:** 10.64898/2026.04.23.26351604

**Authors:** Esha Sheth, Lindsay Case, Fiona Shaw, Nicola Dwyer, John Poland, Yu Wan, Beatriz Larru

## Abstract

**Background:** *Pseudomonas aeruginosa* is a major cause of healthcare-associated infections in paediatric settings, where its persistence in moist environments such as hospital water and wastewater systems poses a particular risk to neonates and immunocompromised children.

**Aim:** The aim of this study was to showcase the long-term survival and transmission of *P. aeruginosa* in a large tertiary children’s hospital in England which is crucial to develop strategies for water-safe care.

**Methods:** Environmental *P. aeruginosa* isolates were collected from taps, sinks, showers, and baths in augmented care areas of a 330-bed tertiary children’s hospital built to NHS water-safety standards. Clinical isolates were classified as invasive (blood, cerebrospinal fluid, and bronchoalveolar lavage) or non-invasive (respiratory, urine, ear, abdominal, and rectal surveillance). Variable number tandem repeat (VNTR) profiles and metadata were extracted from PDF reports, de-identified, deduplicated, and curated using Python and R.

**Findings:** This retrospective study analysed nine-locus VNTR profiles of 457 *P. aeruginosa* isolates submitted to the UK Health Security Agency from a large tertiary children’s hospital, identifying 56 isolate clusters (each with ≥2 isolates), of which 19 (34%) contained at least one invasive isolate. The most persistent cluster (Cluster 1, n=20) spanned from July 2016 to September 2024, containing environmental and clinical (invasive and non-invasive) isolates.

**Conclusion:** These findings demonstrate long-term persistence of certain genotypes and temporal overlap between environmental and clinical isolates, highlighting the difficulty in detecting and eradicating *P. aeruginosa* in hospital water and wastewater systems and reinforcing the need for continuous rigorous water system controls.

## Introduction

In healthcare settings, *Pseudomonas aeruginosa* is an Opportunistic Plumbing Premise Pathogen (OPPP) uniquely suited to colonise and survive in environments linked to water or with high humidity [1,2]. As an aerobic bacterium, *P. aeruginosa* thrives best in the distal parts of the water and wastewater distribution systems, such as taps and sinks where oxygen is readily available. From the periphery of water and wastewater distribution systems, *P. aeruginosa* can be transmitted to patients via direct routes such as contaminated water, aerosols or splashes from water outlets or indirect routes such as contaminated hands or equipment [3]. Several properties enable *P. aeruginosa* to adapt to the hospital environment, including the intrinsic resistance to commonly used disinfectants, largely driven by biofilm formation. These biofilms create a protective matrix that helps bacteria withstand hostile conditions [4].

*P. aeruginosa* is a leading cause of healthcare-associated infections (HCAI) with rapidly growing multi- and pan-antimicrobial resistance [5]. Infections are more likely to occur in patients with weakened immune systems such as those in neonatology, burns or intensive care units (ICUs) and those with underlying conditions like cancer or cystic fibrosis. Outbreaks of *P. aeruginosa* infections linked to water sources in healthcare settings have been frequently reported, mostly in ICUs, oncology, and neonatal wards [3]. *P. aeruginosa* outbreaks are usually slow to recognise, particularly when non-resistant strains are involved, since infections typically manifest as sporadic cases with prolonged infection-free intervals [6]. These outbreaks are also difficult to control despite rigorous implementation of standard infection prevention and control (IPC) measures [7] and may persist over prolonged periods of time with long gaps between patient cases [3,6], making it difficult to link cases and identify environmental reservoirs.

Molecular typing methods, such as variable-number tandem repeat analyses (VNTR), have been shown to enhance IPC by enabling rapid detection of outbreaks and identification of transmission pathways [8]. The UK Health Security Agency (UKHSA) has employed a nine-locus VNTR scheme since 2009 for *P. aeruginosa* typing, generating profiles that are comparable across laboratories and over time for uncovering pathogen transmission and environmental persistence [8].

Understanding the long-term survival and transmission of *P. aeruginosa* in hospitals is crucial to develop strategies for water-safe care. Here, we report a retrospective observational study on the spatiotemporal distribution of *P. aeruginosa* over nine years in a large tertiary children’s hospital in the North West region of England.

## Methods

### Ethics

The study was registered as an audit (Reference 7379) by the Governance and Quality Assurance Department of Alder Hey Children’s Hospital with a waiver of informed consent.

### Clinical setting

Opened in October 2015, the new Alder Hey Children’s hospital is a 330-bed tertiary care centre providing care to over 400,000 children each year. More than 80% of beds are designed as single-bed en-suite rooms. The hospital was built in compliance with NHS England’s Health Building Note 00-10: Part C – Sanitary assemblies (formerly, Health Technical Memorandum 64) and Health Building Note 00-09, with at least one hand-wash sink in each patient room.

### P. aeruginosa isolates

Environmental isolates were collected from various water sources in augmented care clinical areas, specifically taps, sinks, showers, and baths located in wards and operating theatres. Clinical isolates were categorised as possibly invasive or non-invasive based on the sampling site. Invasive isolates were obtained from blood cultures, cerebrospinal fluid (CSF), or bronchoalveolar lavages (BAL). Non-invasive isolates were obtained from all other sites, including upper respiratory samples, urine, ear swabs, and abdominal swabs, as well as rectal surveillance swabs. When transmission between patients and/or patient-associated water sources was suspected, environmental and clinical isolates were sent to UKHSA reference laboratory for VNTR typing at the discretion of Alder Hey’s IPC department.

### Data acquisition

Typing reports generated by UKHSA in the PDF format for referral isolates collected between January 2016 and December 2024 were included in this study [8]. The VNTR typing was conducted using a nine-locus scheme [8], which represented each isolate with a profile of nine integers, each indicating the repeat number of a specific marker locus. A dash in the profile indicates no amplicon was obtained from a specific locus. VNTR profiles and associated metadata (collection and receipt dates, sources) of isolates were extracted from the typing reports and combined into a spreadsheet with custom Python and R scripts. R code was developed to 1) de-identify metadata before analysis, 2) deduplicate VNTR profiles for one isolate per profile per patient/source identifier, 3) impute missing collection dates with the receipt dates at the reference laboratory.

### Data analysis

Isolates were clustered based on 100% identity of the VNTR profiles, and the size of each cluster was defined as the number of isolates sharing the same profile. Single-isolate clusters were excluded from downstream analysis. Remaining clusters were numerically labelled in a descending order of cluster sizes, with Cluster 1 representing the largest cluster, and a distance matrix was generated from VNTR profiles pairwise by counting the number of discrepant loci. A hierarchical clustering dendrogram was constructed in R using the hclust using the complete linkage and visualised alongside metadata with iToL v7 (itol.embl.de) (Supplementary Fig. S1).

## Results and Discussion

### Review of *P. aeruginosa* isolates between 2016 and 2024

The final dataset after deduplication of 28 isolates (25 clinical, 3 environmental) comprised 404 isolates (297 clinical and 107 environmental). A total of 56 clusters with unique VNTR profiles were identified, ranging in size from 20 isolates (Cluster 1) to 2 isolates (Cluster 56) (Supplementary table S1 and Fig. 1). Figure 1 illustrates the temporal distribution of genetically similar VNTR profiles, with 13 clusters containing a mixture of environmental and colonisation isolates and 3 isolates containing a mixture of environmental, possible invasive and colonisation isolates. The longest detection span was observed in Cluster 1 (n = 20), with *P. aeruginosa* first detected in July 2016 and last detected in September 2024. This cluster included 17 environmental isolates (8 from critical care outlets, 6 from environmental outlets, 1 from a theatre sink and 6 from unknown sources) and 3 clinical isolates, of which one (from a blood culture) was classified as invasive. Our results show persistence of *P. aeruginosa* Clusters 1, 3, 4, and 6 from 2 to 8 years for which the corresponding VNTR profiles are listed in Supplementary table S1.

**Figure 1.**
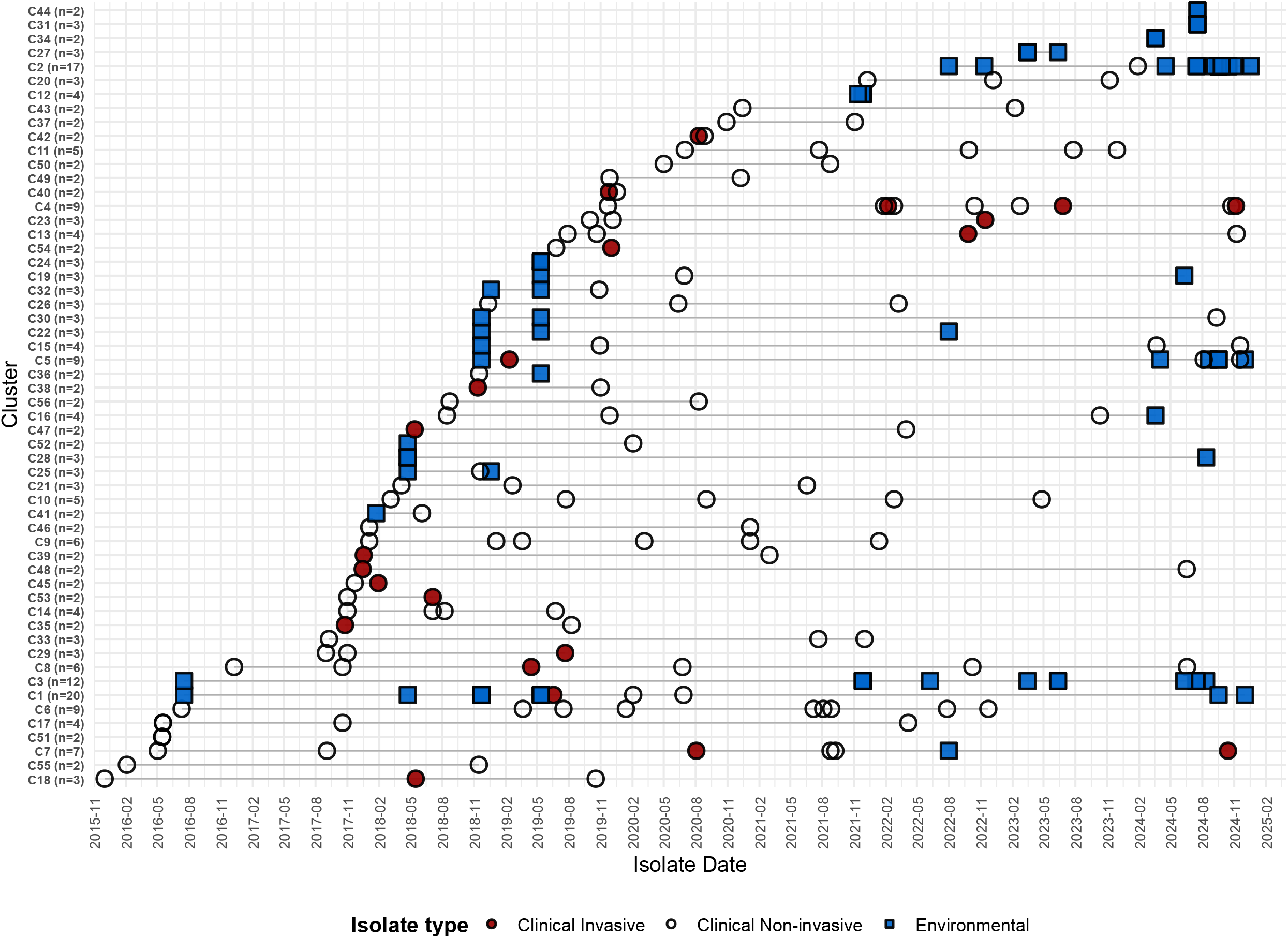
Temporal graph of non-singleton clusters of *P. aeruginosa* isolates. Horizontal lines represent the period from first to last detection of each VNTR profile. Cluster labels, such as “C1 (n=20)”, show the ranks and isolate numbers.

Persistence of microbial communities including *P. aeruginosa* in newly built hospitals for extended periods has previously been described and novel technologies, such as deep shotgun metagenomics or whole-genome sequencing (WGS), have advanced our understanding of the hospitals microbiome as reservoirs of antimicrobial resistance (AMR) [9]. Water and wastewater systems where bacteria can form biofilms and persist for years interacting with antibiotic residues and heavy metals are critical reservoirs of AMR and act as persistent source of nosocomial infections. Our study strengthens the evidence of cross-transmission of *P. aeruginosa* between patients and hospital environments and underscores the need to establish effective methods of preventing transmission and eradicating *P. aeruginosa* from plumbing systems.

In our study, we divided *P. aeruginosa* isolates obtained from clinical specimens as representing invasive infection or colonisation, depending on the anatomical site where the specimen was obtained. We only considered isolates from blood, CSF, or BAL as invasive, due to limited clinical data. Despite this strict criterion, out of the 56 clusters we identified, 19 (34%) contained at least one invasive isolate (Fig. 2). Infection cases were distributed across clusters with detection spans ranging from 3 months (Cluster 15 [n=2]; November 2017 to February 2018) to over 8 years (Clusters 1 [n=20], 3 [n=12], and 51 [n=7]; 2016 to 2024). These observations highlight the potential for long-term cross-contamination of certain *P. aeruginosa* genotypes in the hospital environment, with environmental detections preceding clinical cases in 9 clusters (Fig. 1). Of these, 2 clusters (Clusters 1 [n=20] and 3 [n=9]) involved invasive cases (Fig. 2), while the remaining 7 clusters involved only colonising or non-invasive cases. Transmission of *P. aeruginosa* from water outlets causing colonization/infection in patients is frequently not recognised outside outbreak settings or when caused by non-resistant strains. Our study shows that colonisation of water outlets with *P. aeruginosa* acts as a persistent source of infections for patients and highlights the urgent need to maintain control measures to reduce *P. aeruginosa* colonisation in hospital water.

**Figure 2.**
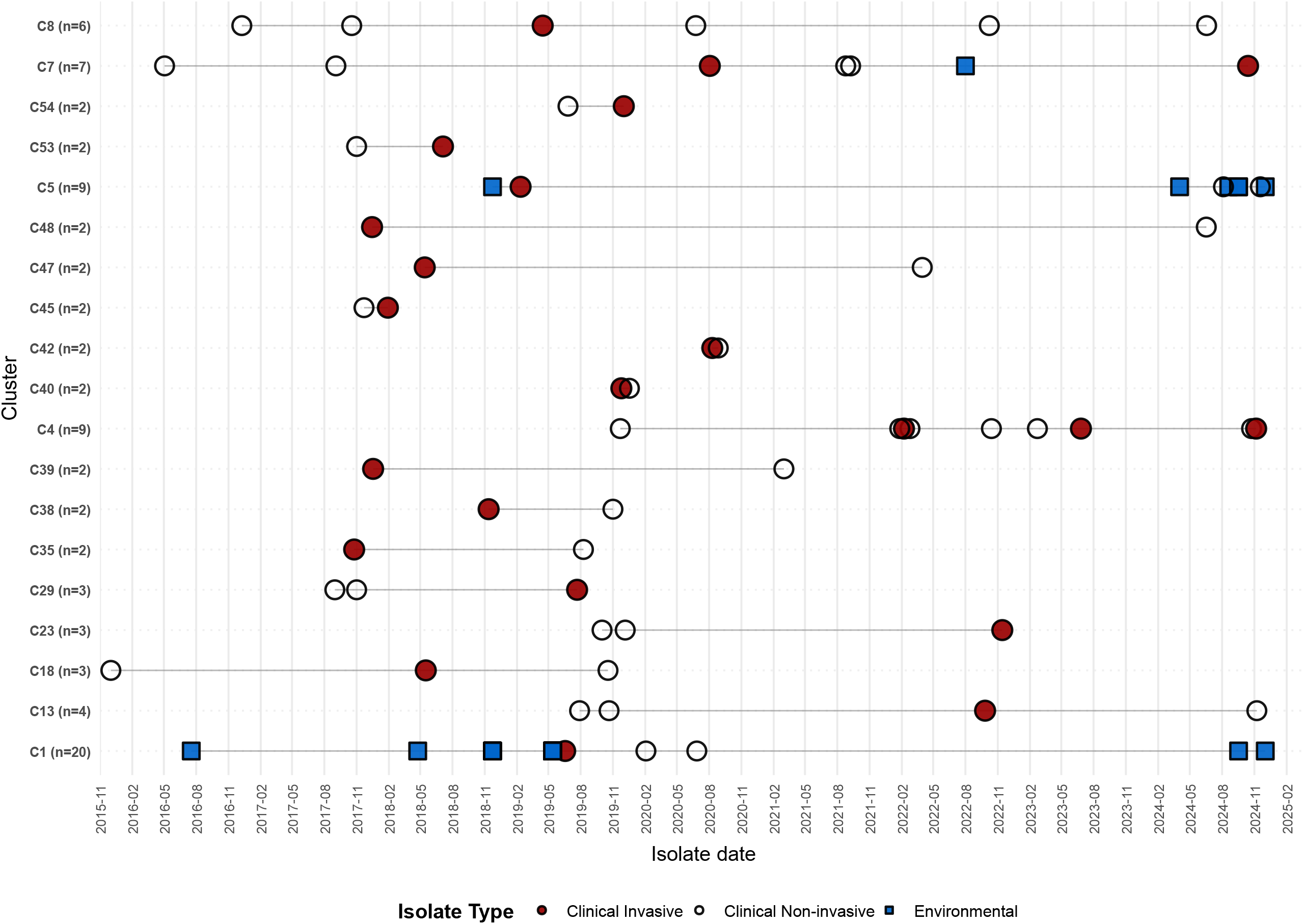
Temporal graph of *P. aeruginosa* VNTR clusters containing at least one invasive isolate. Horizontal lines represent the period from first to last detection of each VNTR profile. Cluster labels, such as “C1 (n=20)”, show the ranks and isolate numbers.

### Limitations

There were notably several limitations in our study. The retrospective design of our study lacked a systematic typing to all *P. aeruginosa* detected isolates and variable sampling intervals during the study period may have led to underestimation of some isolates in the temporal trends. We might have underestimated invasive cases because while the nine-locus VNTR scheme is effective for identifying closely related strains, it has lower discriminatory power to that of the WGS, which has the capacity to detect AMR genes, virulence factors, and other clinically relevant genetic features [2,10]. Without the WGS, we cannot be certain about the transmission pathways. However, the detection of 9 clusters in environment samples prior to clinical cases may be used to support the hypothesis for the transmission of *P. aeruginosa* from environmental sources to patients. Another important limitation was the restricted spatial resolution of the metadata accompanying environmental samples. Precise sampling locations were not available for 361 (89%) isolates, which prevented the interpretation of environmental dispersion patterns in our setting.

## Conclusion

The long detection spans and overlap between environmental and patient isolates in several clusters provide suggestive evidence of healthcare-associated transmission of *P. aeruginosa* linked to water and wastewater systems and prolonged persistence of *P. aeruginosa* contamination in healthcare settings. These findings support continued emphasis on ensuring water safe care in paediatric hospitals.

## Data Availability

All data produced in the present work are contained in the manuscript and supplementary table.

## CRediT authorship contribution statement

**Esha Sheth**: Investigation, Data curation, Formal analysis, Software, Visualisation, Writing - Original draft, Writing - Review & Editing, Funding acquisition. **Lindsay Case**: Project administration, Investigation, Data curation, Writing - Review & Editing. **Fiona Shaw**: Investigation, Writing - Review & Editing. **Nicola Dwyer**: Investigation, Data curation, Writing - Review & Editing. **John Poland**: Investigation, Data curation, Writing - Review & Editing. **Yu Wan**: Methodology, Software, Investigation, Data curation, Formal analysis, Writing - Original draft, Writing - Review & Editing, Supervision, Project administration, Funding acquisition. **Beatriz Larru Martinez**: Conceptualization, Methodology, Investigation, Data curation, Writing - Original draft, Writing - Review & Editing, Supervision, Project administration, Funding acquisition.

## Acknowledgements

We thank Clare Kahuma, Henry Mutegeki, and Ellon Twinomuhwezi at the CAMO-Net Uganda Hub and Victoria Winters (CAMO-Net Liverpool site) at the CAMO-Net UK Hub for supporting ES’s fellowship. We also thank the microbiology laboratory staff at Alder Hey Children’s NHS Foundation Trust, for their expertise in sample collection and microbial identification.

## Code availability

Computer scripts for data extraction and analysis are available online at github.com/esheth98/vntr-analysis.

## AI declaration

During the preparation of this work the authors used Microsoft Copilot in order to refine and troubleshoot R code for data analysis. After using this tool, the authors reviewed and edited the content as needed and take full responsibility for the content of the published article.

