## Supplementary figure S1 for "Retrospective analysis of clinical and environmental genotyping reveals persistence of *Pseudomonas aeruginosa* in the water system of a large tertiary children’s hospital in England"

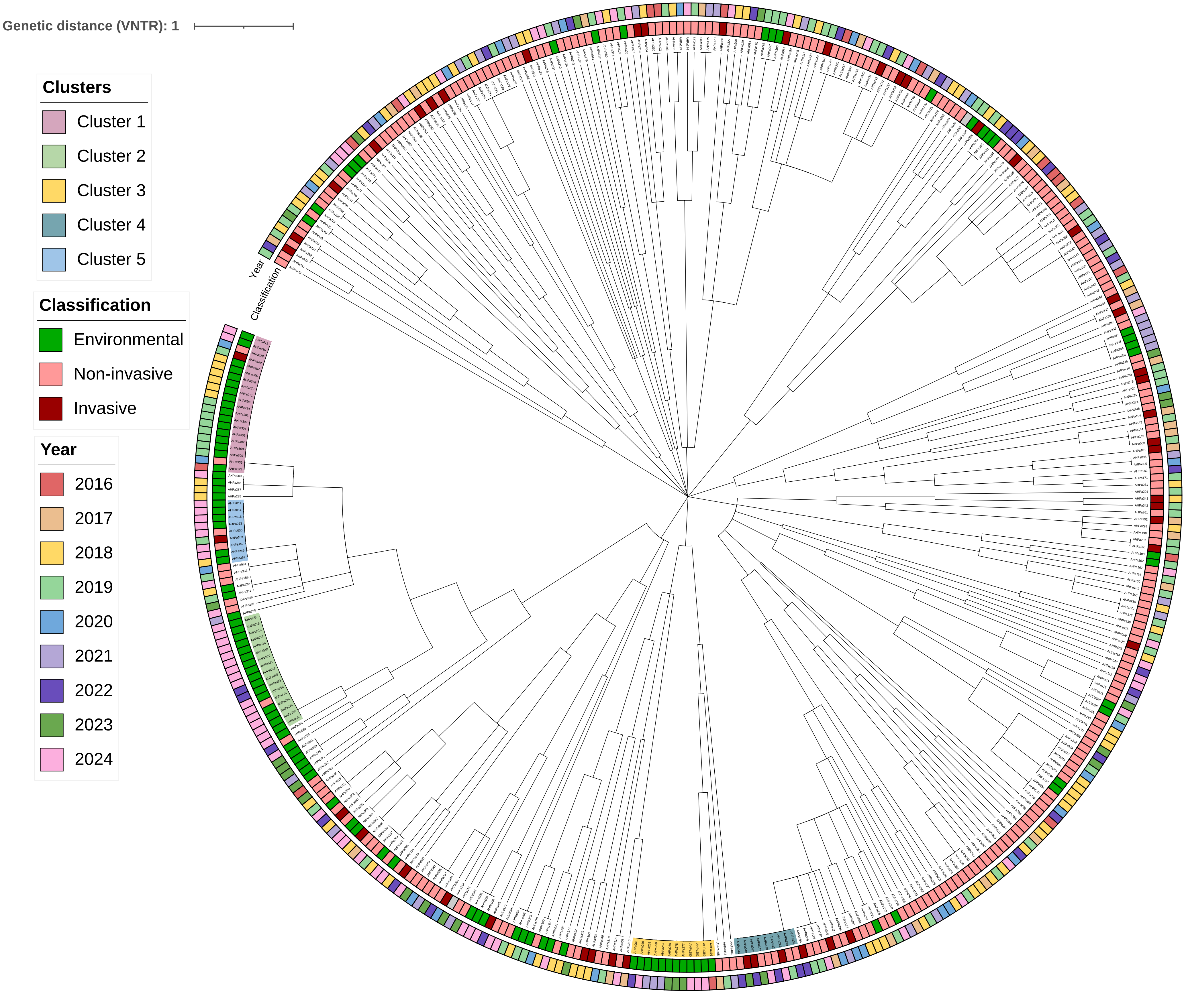


Supplementary Figure S1: Phylogenetic tree of 404 *Pseudomonas aeruginosa* isolates based on VNTR profiles (9 loci). Isolates are labelled with unique AHPa identifiers. The inner colour strip indicates source and isolation class (dark red = possible clinical invasive; light red = clinical non-invasive; green = environmental). The outer colour strip indicates year of collection (2016 - 2024). Clusters 1 to 5 of identical VNTR profiles are labelled.
